# Assessment of Dietary Pattern Among Medical Students of Ebonyi State University

**DOI:** 10.1101/2025.10.13.25337877

**Authors:** Ajana Ikenna Daniel, Igbojimba Benjamin Kelechi, Ukpai Amarachukwu Ejitu

**Affiliations:** Griggs Specialist Hospital,Lagos; Bishop Obot Memorial Hospital, Ankpa; Safeway Hospital,Lagos

**Keywords:** Assessment, Dietary pattern, medical students, Ebonyi, Nigeria

## Abstract

**Background:** Poor eating habits are a major public health concern among young adults who experience transition into university life during which they are exposed to stress and lack of time. This study was designed to assess dietary patterns among medical students of Ebonyi State University, Southeastern Nigeria.

**Methodology:** The study was a descriptive cross-sectional study carried out with systematic random probability sampling for consenting medical students of Ebonyi State University. Data were collected using a structured, self-administered questionnaire. Data entry and analysis were done using IBM Statistical Package for the Social Sciences (SPSS) statistical package version 25.

**Result:** The majority of the students, 228 (55.2%), aged between 20 and 24 years, and they were predominantly 231 (55.9%) females. Almost all,401 (97.1%) of the students were single, and 400 (96.9%) were Christians. On their monthly allowances, 149 (36.1%) and 127 (30.8%) receive less than 10,000 naira and between 10,000 and 20,000 naira, respectively. There is a significant association between the religion of the participants and the determinants of their dietary pattern (t=2.486; P=0.013<0.05).Again,there is a significant association between the level of study of the participants and the determinants of their dietary pattern (F=2.702; P=0.030<0.05). Also, there is a significant association between the monthly allowances received by the students and the determinants of their dietary pattern (F=3.705; P=0.006<0.05).

**Conclusion:** Our findings show that the majority of the medical students have poor attitudes towards diet and low/poor dietary diversity. Also,majority of the students use the selected food 2 to 3 days a week in the past 12 months. Time constraints and proximity of the needed food are the two major determinants of dietary pattern among the medical students. Thus, nutritional intervfentions should be implemented at an individual, family, community, and national levels.

## INTRODUCTION

### 1.0 BACKGROUND

Dietary pattern (DP) is the general profile of food and nutrient consumption which is characterized on the basis of the usual eating habits. The analysis of dietary patterns gives a more comprehensive impression of the food consumption habits within a population. It may be better at predicting the risk of diseases than the analysis of isolated nutrients or foods because the joint effect of various nutrients involved would be better identified.^1^

Eating is vital in life and a major determinant of health hence it is important to study this subject from its different perspectives. Most studies in eating and nutrition has focused on physiological limited knowledge. It is for this reason that cultural, psychological and social approach is necessary. Different attitudes towards food may have an effect on overall health but if dissociated from their relevant social environment, it is believed to produce only differences in non-communicable diseases.^1^

Nutritional status is the sum total of an individual’s anthropometric indices as influenced by intake and utilization of nutrients, which is determined from information obtained by physical, biochemical, and dietary studies result of interrelated factors influenced by quality and quantity of food consumed and the physical health of the individual.^2^ The main factors for eating are hunger and satiety, however what we choose to eat is not determined exclusively by the physiological or nutritional needs. On the other note, an adolescent nutritional status has important implications for his health, development of several chronic diseases, and plays a key role in breaking the cycle of malnutrition.^2^

The transition from adolescence to adulthood is an important period for establishing behavioral patterns that affect long-term health and chronic disease risk. University students seem to be the most affected by this nutritional transition.

Patterns of nutritional behaviors adopted in childhood and adolescents are mostly continued in adult life and increase the risk of development of many chronic diseases.^3^

Diet in childhood and adolescents have public health implications due to evidence relating poor nutrition in childhood to subsequent obesity and elevated risks for type2 diabetes, metabolic syndrome, and cardiovascular diseases.

Food intake patterns and overweight are associated with multiple rapid complications such as heart disease, hypertension stroke, cancer, dental caries, asthma, and some other psychological disorder like depression.^4^

Poor eating habits is a major public health concern among young adults who experience transition into university life during which they are exposed to stress and lack of time.

For the first time in their lives, university students experience independence and freedom from their parents’ supervision. In this new environment, different settings may cause questioning of the parental values which are learned from childhood. Rozmus et al suggests that different views and dietary lifestyles are moulded and shaped by this new environmental, social, personal and financial stressors in these surroundings.^5^

This poses a barrier against adoption of healthy behaviours and university students seem to be the most affected by this nutrition transition. Studies from developed countries have shown that young adults living away from home to attend college experience numerous health-related behavioural changes, including the adoption of unhealthy dietary habits.^4^

These behaviours are mostly attributed to drastic changes in the environment, available resources, frequent exposure to unhealthy foods and habits leading to higher consumption of high caloric snacks, fast foods, and lower consumption of fruits and vegetables, i.e replacing their consumption of nutrient-dense foods with energy-dense nutrient-poor foods. In addition to that, skipping meals may also become more frequent.

It has been assumed that medical students would practice healthy dietary habits compared to non medical students, on down note, some studies have found otherwise. A study done in China have revealed that medical students exhibited early risk factors for chronic diseases due to poor eating habits. It is alarming one can say how medical students being the most up to date on latest health practices are the least active in implementing that knowledge in their lifestyle.^5^ The feeding lifestyle of university students is associated with higher consumption of fried food, meats, sugar salt, convenience food (frozen, ready to eat foods), cereals, bread, dairy products, soda beverages, snacks, and less fruits and vegetables.^6^

### 1.1 PROBLEM STATEMENT

Dietary patterns of young adults have been widely studied and reported in the literature, as being associated with obesity, frequent snacking and meal skipping.

Type II diabetes particularly is on the rise with a national prevalence of 4.42%. This is attributed to dietary transition from traditional diet to western diet which is characterized with high calroric intake.

Out of 800 students of a private University in Southern Nigeria; 196(24.5%) had a medium of dietary diversity. 112(14.0%) were overweight and 76(9.6%) were obese.^5^

Diet plays an important role in the development of type II diabetes so it becomes pertinent that one is equipped with knowledge of which foods to eat in order to balance sugar levels.

Diabetic patients tend to have magnesium inadequacy and also loose it in urine. This deteriorates their health condition and leads to diabetic-related complications. Consumption of whole grains, fruits and vegetables provides the body with magnesium. Magnesium is important in regulating their blood glucose levels and also prevent/reduce incidence of diabetes-related complications.^5^

Furthermore a study conducted among international students in the United States revealed unhealthy dietary pattern throughout the university life and the probability of continuity has been confirmed to be greater by previous researchers.

The feeding lifestyle of University students is associated with higher consumption of fried food, meats, sugar salt, convenlence food (frozen, ready to eat foods), cereals, bread, diary products, soda beverages, snacks, and less fruits and vegetables.

Finally, no research on dietary pattern have been carried out in eastern part of Nigeria.

### 1.2 JUSTIFICATION

The Management of Nigerian institutions have not really placed much concern on the level of knowledge Nigerian students have on good nutrition. Studies have shown that the level of education can influence dietary behaviour during adulthood. Some common unhealthy eating patterns among young adults in the university are products of many factors which could range from ignorance, lack, inability to adapt, laziness and environmental factors. For instance, awareness on health benefits of consuming whole grains, fruits and vegetables improves healthy food choices that help regulate the blood glucose levels.

According to Convey, habits are developed at the intercept of knowledge, skill and desire (attitude). Studies have identified significant barriers that prevent health workers from dietary support as lack of Nutrition knowledge and confidence on the part of the provider. Therefore tertiary institution should critically appraise the training given to students undergoing medical training in health profession.^7^

With up to date knowledge of good dietary pattern, the growing youths that dominate high institution although far from home can still live a healthy lifestyle in school.

Furthermore, the research on assessing the Dietary Pattern among undergraduate students has been done in some developed Nations but none has been done in eastern part of Nigeria. Although it has been carried out in the southern part of the country.

### 1.3 RESEARCH QUESTIONS

1. What is the attitude of medical students towards diet?
2. What is the pattern of diet among the medical students?
3. What is the dietary diversity score of medical students?
4. What are the factors that determine the nutrition pattern of medical students?

### 1.4 GENERAL AND SPECIFIC OBJECTIVES

#### 1.4.1 GENERAL OBJECTIVE

To assess the prevalence of food consumedby the medical students in Ebonyi State University.

#### 1.4.2 SPECIFIC OBJECTIVES

1. To assess the attitude of medical students towards diet.
2. To assess the pattern of diet among the medical students.
3. To ascertain the dietary diversity score of medical students.
4. To ascertain the factors that determine the dietary pattern of the medical students.

## CHAPTER TWO

### 2.0 LITERATURE REVIEW

The term “diet” generally refers to the foods people habitually consume or are on a set way of eating for medical reasons.^8^

Dietary pattern is the quantities, proportions, variety, or combination of different foods and drinks in diet, and the frequency with which they are habitually consumed.^9^Some of the dietary patterns include:

#### Vegetarian/low caloric dietary pattern

characterized mainly by consumption of plant-based food while avoiding ‘western” food, composite dishes, and bread; **a mixed or Mediterranean dietary pattern** characterized by high consumption of plant-based food, followed by composite dishes, bread, and a low consumption of western type food; and finally, **a westernized dietary pattern** characterized by high consumption of white bread and western food, and a strong avoidance of plant food and composite dishes.^10^

The complex relationship of nutrients in our diets can help provide evidence for positive health outcomes with changes to our total diet not just one aspect of it. It also provides practical information for the public as people choose to eat foods, not nutrients.

So as the times are changing, and we move away from writing to typing on ipads and tablets, nutrition is also changing. Nutrition is steering away from single nutrients but now focusing on what foods, how the nutrients in these foods interplay with each other, the variety of foods and the quantity of foods we consume over days, weeks, months and years and how this might affect our health outcomes.^11^ However, nutritional adequacy as well as micronutrient malnutrition (also called ‘hidden hunger’) in an individual can be validly indicated by the use of Dietary Diversity Score (DDS). According to World Health Organization (WHO), Dietary diversity Score is mostly determined by counting the number of selected food groups of about nine, consumed by individuals or a household over a reference period, which usually ranges 1-3days, and in some cases 7days. These nine food groups include: 1. Starchy staples (cereals, roots, tubers), 2. Vitamin A rich fruits and vegetables, 3. Other fruits, 4. Other vegetables, 5. Legumes and nuts, 6. Fats and oils, 7. Meat, poultry, fish, 8. Milk and milk products, 9. Eggs.

### 2.1 MAJOR DETERMINANTS OF DIETARY PATTERN

The different stages of life, in particular work or study-related, can produce profound changes in eating habits. The start of university education is an important time in the life of an individual, since it often represents a period of greater responsibility for food choices and health.^12^

The most common factors influencing eating habits include: Biological determinant such as sex, hunger, appetite and taste, even though what we eat is not determined solely by physiological or nutritional needs.

Economical determinants such as costs and financial resources. Physical determinants such as skills (e.g cooking), time, living arrangements, availability of convenience and fast meals.

Social determinants such as culture, religion, family, peers. Psychological determinants such as mood, stress, anxiety and guilt.

#### Biological determinants of dietary pattern

Our physiological needs provide the first line determinant of eating habit. Energy and nutrient are required by humans to survive, this is why humans respond to hunger and satiety.^13^ The lateral hypothalamic area and the ventro-medial hypothalamus of the central nervous system is involved in the control of the balance between hunger and satiety respectively.^14^ The macro-nutrients i.e carbohydrates, proteins and fats generate sense of fullness (satiety) of varying degree. However, evidence has proved that fat, carbohydrate, and protein has the lowest, intermediate and highest satiating capacity respectively. Thus it can be inferred that the degree of satiety derived from a diet is inversely proportional to energy density of the diet. Therefore, high energy food can also lead to “passive overconsumption” in which there is excessive and unintentional caloric intake without a commensurate consumption of additional bulk.^15–17^ Sensory properties of food such as taste, smell, texture, and appearance influences the pleasure someone experiences when eating a particular food; this is known as palatability. However, palatability and taste in turn influence eating habits.

#### Economic and physical determinants of dietary pattern

It is unarguable that the cost of food is primary determinant of eating habit. Low-income groups (students) have a great tendency to consume unhealthy diets with infinitesimal intake of fruit and vegetables. By implication access to more money is not automatically equal to a better healthy diet but the range of foods from which one can choose increases. Availability of convenience and fast meals, an important physical factor influencing eating habit, is dependent mainly on geographical location. Time constraints will prevent individuals from adopting healthy diets, especially the students that live alone who choose convenience foods. This is also a key in student’s dietary pattern.^18^

#### Social determinants of dietary pattern

Social and cultural circumstances forms what people eat. Population study show that there are clear differences in social classes with regards to food and nutrient habit. Cultural influences lead to the difference in the habitual consumption of certain diets and traditions of preparation, and in particular cases lead to restrictions such as exclusion of meat and milk from the diet.^18^

Social influences on dietary habit refer to the impact that one or more persons have on the eating behaviour of other, either direct (buying food) or indirect (learn from peer’s behaviour) either conscious (transfer of beliefs) or subconscious. However, quantifying the social influences on dietary habit is herculean because the influences that people have on the eating behaviour of others are not limited to one type and people are not necessarily aware of social influences that are exerted on their eating behaviour.^18^

The family is strongly recognized as being significant in dietary pattern. Research shows that shaping of feeding habits takes place in the home. Because family and friends can be a source of motivation in making and sustaining dietary change. Adopting dietary strategies which are acceptable to them may benefit the individual whilst also having an effect on the eating habits of others.

#### Psychological determinant of dietary pattern

Psychological stress is a common feature of modern life and can modify eating habits. Thus, mood and stress can influence the eating habit and possibly short and long term responses to dietary intervention.^19^

### 2.2 THE IMPACT OF DIETARY PATTERN ON HEALTH

The role of healthy eating in the prevention of chronic and infectious disease has been well documented. A balanced diet and consumption of food prepared in accordance with good practices are factors that contribute to maintaining a healthy lifestyle.^20^ The Mediterranean diet is widely recognized as satisfying the requirements of healthy nutrition. Originally, this diet may be pictured as a pyramid which represents food types and frequency of consumption: (a) high intake of vegetable, pulses, fruits, dietary fibers and cereals; (b) medium-high intake of fish; (c) olive oil as the principal fat; (d) medium-low intake of dairy products; (e) low intake of meat and saturated fat (f) moderate consumption of alcohol. Moreover, departure from the ideal model of the Mediterranean diet may increase risk of a spectrum of disease conditions.^21–22^ Some classes of food with their health implications are:

#### Carbohydrate

It has been reported that high intake of carbohydrate is linked to obesity, diabetes mellitus, cancer and emotional problems. Fermentable sugars have also been implicated in dental caries and gingivitis while sugar alcohols and artificial sweetners are not fermentable sugars and do not cause dental problems.^23^

Fats and oil: It has been proved that saturated fats (fatty meat, butter palm oil, coconut oil) is linked to cardiovascular diseases such as high blood pressure, high cholesterol, ischaemic stroke, heart attack and arrhythmia while unsaturated fats (olive oil, vegetable oil, soy bean, avocado, fish and nuts) have been linked to optimal cardiovascular functions and transmission of brain signals. Also potatoes, yams, meats, fishes which are rampantly fried either with vegetable oil or red oil at a very high temperature have been proven to be carcinogenic. This is due to the release of polycyclic aromatic hydrocarbons and heterocyclic amines (carcinogens) from heated oils.^24^

Dietary Fibers: Low intake of dietary fibers have been associated with diverticulitis and diverticulosis while high intake of dietary fibers have been associated with low blood cholesterol, low incidence of colon cancer, weight reduction, low risk of type 2 diabetes and low risk of cardiovascular diseases. On the other hand, consumption of food away from home is often associated with food-born disease outbreaks.^25^

#### Vitamins

It has been documented that low intake of foods (fruits and vegetables) rich in vitamin B complex is associated with impaired energy metabolism, anemia, pallegra and beriberi. The inhibitory role of alcohol on the absorption of vitamin BI (thiamine) has been implicated in wernicke encephalopathy and ultimately, korsakoff’s psychosis.^26^

### 2.3 THE IMPACT OF DIETARY PATTERN ON ACADEMIC PERFORMANCE

Research has proven that healthy dietary patterns of students showed a positive association with their academic performance. However, unhealthy dietary patterns such as skipping breakfast and regular meals, excessive intake of fast foods/meals and soft drinks together with an inadequate intake of fruits, vegetables and fish may have serious consequences related to poor academic performance among students. Breakfast is an important factor which improves the cognitive and academic performances by enhancing postprandial memory functions. Intake of fresh fruits and vegetables is linked to better mental health such as positive mood, greater happiness, socio-emotional flourishing and overall wellbeing.

High intake of foods (fish, walnut) rich in w-3 PUFA help to maintain the balance between w-3 and w-6 at a normal ratio of 1:1 to 1.5. This is linked with higher school grades, whereas fast foods/meals were observed to be linked with poor academic performance among students. This is due to the fact that food processing industries and fast food vendors uses hydrogenated oil in food preparation. In hydrogenated oil, there is imbalance in the ratio of w-3 PUFA to w-6 PUFA. This imbalance is associated with neurodegenerative diseases like dementia which destroys memory and thinking skills.

The B-vitamins, particularly folate, vitamin B12, and vitamin B6, are widely believed to be protective against Alzheimer’s disease and related cognitive decline. The primary features of Korsakoff’s psychosis (thiamine deficiency) are loss of memory, confabulation, and hallucination. This rare syndrome is most commonly the result of chronic alcoholism. Even if there is adequate nutritional intake, chronic alcoholism affects thiamine metabolism.^27^

## CHAPTER THREE METHODOLOGY

### 3.1 STUDY AREA

The study area was Ebonyi State University in Abakaliki LGA of Ebonyi State, Nigeria .Ebonyi State University has a total number of 19,753 students. The study area consist of about eleven(11) faculties. This study location consist of a wide range students of different socio-economic classes, families, cultures, and religions. It is located in the capital and largest city of Ebonyi State which is a south-eastern state of Nigeria, populated primarily by Igbos(95%). This study area is situated between longitudes and latitudes (6.264°N) (8.013°E) and has a last known population of 2,880,383 (year 2016). The state shares a border with Benue State to the North, Enugu State to the West, Imo State and Abia State to the south and Cross River State to the East. The study area also consists of many state and local government funded primary and secondary school, private schools and three universities. Other commercial cities are Afikpo, Ishieke, Ohaozara, and many others.

### 3.2 STUDY LOCATION

The study location consists of about three faculties including faculty of basic medical sciences, faculty of basic clinical sciences, and faculty of clinical medicine.

### 3.3 STUDY POPULATION

The study population was about 700 students from a wide range of different socio-economic classes, families, cultures, and religions who have different dietary habits.

### 3.4 INCLUSION CRITERIA

The selection criteria includes those medical students that were present in school as at the time of administration of the questionnaire.

### 3.5 EXCLUSION CRITERIA

The medical students that did not give consent in answering the questionnaire.

### 3.6 STUDY DESIGN

The study design is a descriptive cross-sectional study. The instrument for getting the information was the questionnaire using lickert scale.

### 3.7 DETERMINATION OF SAMPLE SIZE

The minimum sample size usedfor this study was calculated using the sample size formula given below:

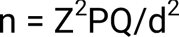

where n = desired sample size

Z = standard normal deviation usually set at 1.96

P= prevalence set at 50% = 0.5

Q = 1-P = 0.5

d = degree of accuracy desired, usually set at 0.05 (i.e 95% accuracy is desired) = 100-95 = 5% = 0.05.

apply the above formula

n = [(1.96)^2^ x 0.5 x0.5]/0.05^2^

n =[3.8416 x 0.5×0.5/0.0025

n = 384.16 (approximately 384)

adjustment for none-response: n +10% of sample size = 384+38 = 422

### 3.8 SAMPLING METHOD

The systematic randomprobability sampling method was used and participants selected were consenting medical students who satisfied the selection criteria.

### 3.9 DATA COLLECTION

Data was collected from participants using structured, self-administered, pretested questionnaire. This was preceded by due explanation of the purpose of the study, reassurance about the confidentiality and obtaining consent.

### 4.0 DATA ANALYSIS

The data was analysed using the IBM SPSS version 25 statistical package for sciences and presented in simple frequency tables and calculated in proportions and percentages.

### 4.1 ETHICAL CONSIDERATIONS

Permission for the study was obtained from the ethics and research committee of Alex Ekwueme Federal University Teaching Hospital, Abakaliki. A written consent was obtained from each participant before administering the questionnaire. Each respondent was assured of maintenance of their privacy and confidentiality by leaving out their names to ensure anonymity. Those who were not willing were left out.

### 4.2 LIMITATIONS OF THE STUDY

Inability to carry out a total population study.

## CHAPTER FOUR RESULT

The results obtained from data analysis were presented in this chapter. Four hundred and twenty-two (422) copies of questionnaires were administered; four hundred and thirteen(413) were returned, properly filled and fit for analysis. Therefore, the return rate was 97.9% which is a good return rate for a survey. The mean and standard deviation of the age of the students was 22.25±3.98 years. The result of the analysis was presented with tables.

The result in table 1 shows the demographic characteristics of the students. The majority of the students 228 (55.2%) aged between 20 – 24years, and they were predominantly females231 (55.9%). Almost all, 401 (97.1%) of the students were single and 400 (96.9%) were Christians. On their ethnicity, the majority 392 (94.9%) were Igbos. A greater proportion of them, (32.4%) were in 200Level of study, while a minor proportion (8.5%) were in their 600Level. On their monthly allowances, 149 (36.1%) and 127 (30.8%) receive less than 10,000naira and between 10,000 to 20,000 naira, respectively.

**Table 1:** Socio-demographic characteristics of the Students N = 413.

| **Characteristics** | **Frequency** | **Percentage** |
| --- | --- | --- |
| **Age (Years)** |  |  |
| 15-19 | 95 | 23.0 |
| 20-24 | 228 | 55.2 |
| 25-29 | 64 | 15.5 |
| 30 & above | 26 | 6.3 |
| **Gender** |  |  |
| Male | 182 | 44.1 |
| Female | 231 | 55.9 |
| **Marital Status** |  |  |
| Single | 401 | 97.1 |
| Married | 12 | 2.9 |
| **Religion** |  |  |
| Christian | 400 | 96.9 |
| Others (Islam, Atheist, Traditional) | 13 | 3.1 |
| **Ethnicity** |  |  |
| Igbo | 392 | 94.9 |
| Others (Yoruba, Hausa) | 21 | 5.1 |
| **Level of Study** |  |  |
| 200L | 134 | 32.4 |
| 300L | 88 | 21.3 |
| 400L | 84 | 20.4 |
| 500L | 72 | 17.4 |

| Characteristics | Frequency | Percentage |
| --- | --- | --- |
| 600L | 35 | 8.5 |
| **Monthly Allowance (Naira)** |  |  |
| <10,000 | 149 | 36.1 |
| 10,000-20,000 | 127 | 30.8 |
| 21,000-30,000 | 44 | 10.7 |
| 31,000-40,000 | 61 | 14.7 |
| 41,000-50,000 | 32 | 7.7 |

### On the attitude of students towards their diet as presented in table 2

**a** greater proportion (35.1%) of the students take about half of the responsibilities for food shopping. Almost half, 187 (45.3%) take half of the responsibilitiesfor planning for meals. Greater proportion 186 (45.0%) take most of all the responsibilities for preparing meals.

**Table 2:** Attitude towards Diet Among the students.

| **Attitudes** | **Little or**<br>**none (%)** | **About**<br>**half (%)** | **Most of**<br>**all (%)** |
| --- | --- | --- | --- |
| Responsibilities for food shopping | 135 (32.7) | 145<br>(35.1) | 133 (32.2) |
| Responsibilities for Planning meals | 94 (22.8) | 187<br>(45.2) | 132 (32.0) |
| Responsibilities for preparing meals | 96 (23.2) | 131<br>(31.7) | 186 (45.0) |

### Table 3 shows the pattern of diet among the Students

The student take their breakfast in a typical week, 334 (80.9%) and 36 (8.7%) take their breakfast at home and outside, respectively. Out of the 370 that take their breakfast either at home or outside, the majority 123 (33.2%) and 136 (36.8%) oftentimes take their breakfast between 4-5days/week and 6-7days/week, respectively.

**Table 3:**
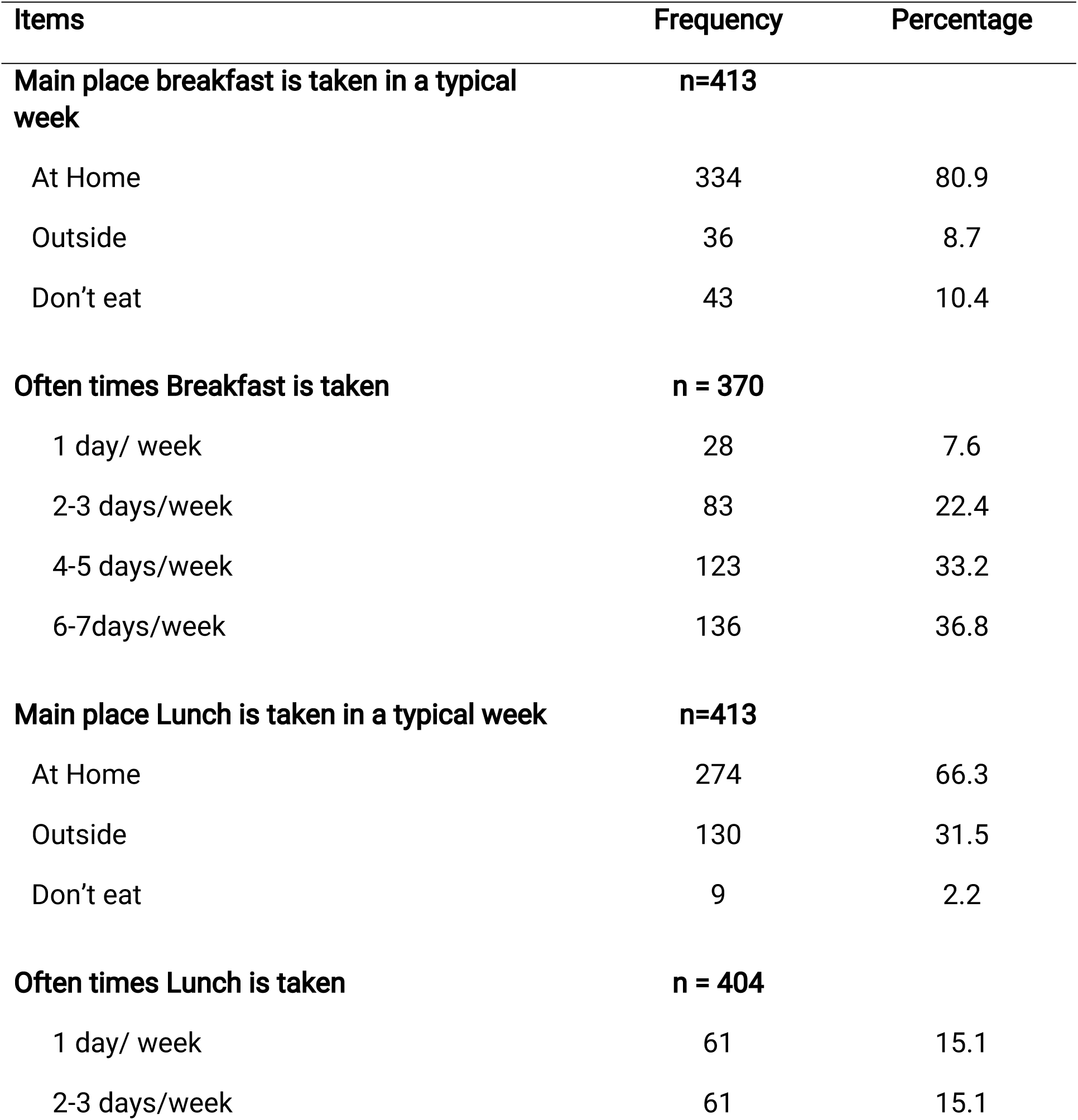
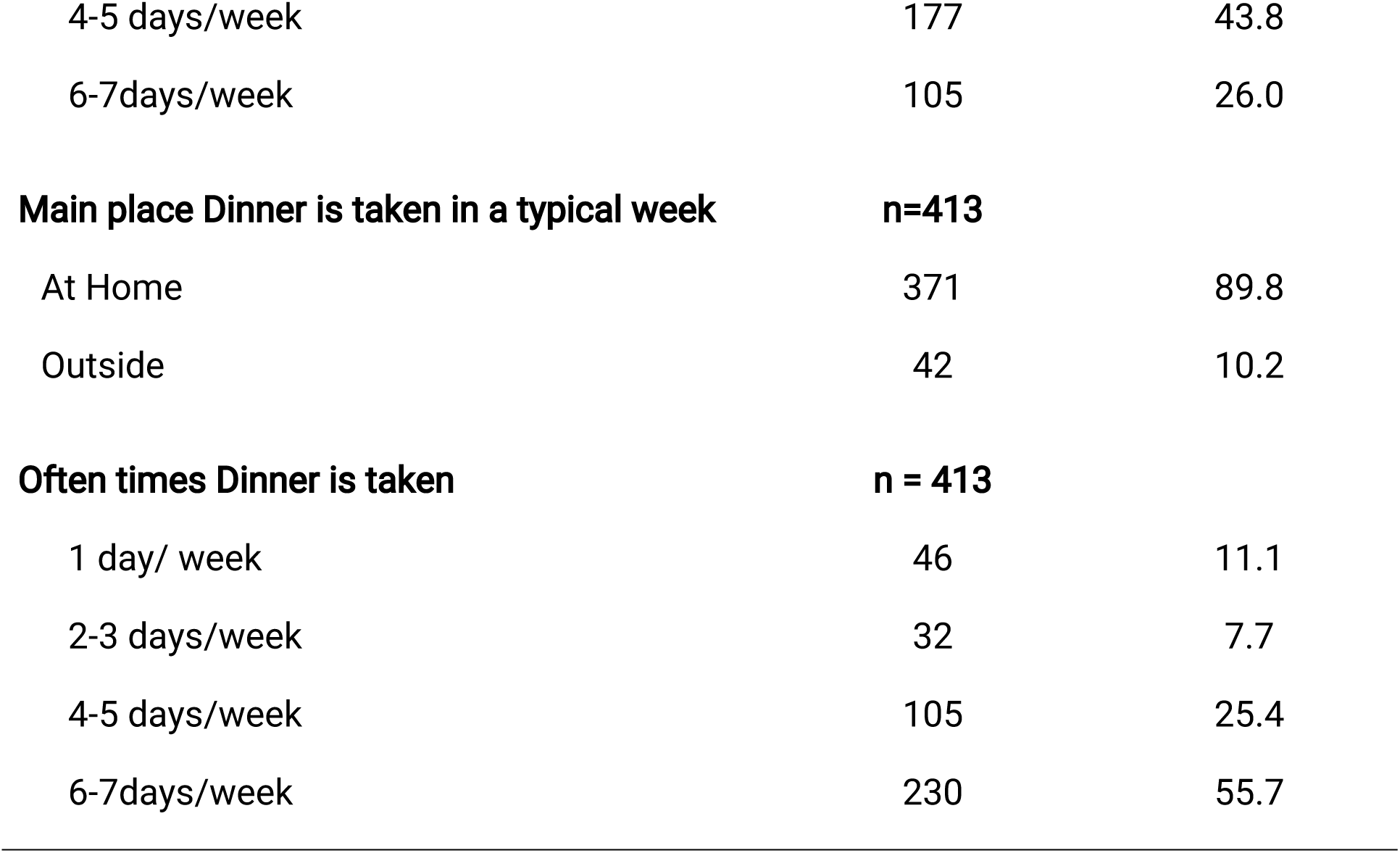
Pattern of Diet among the Students.

On the main place, the students take their lunch in a typical week, 274 (66.3%) and 130 (31.5%) take their lunch at home and outside, respectively. Out of the 404 that take their lunch either at home or outside, the majority 177 (43.8%) and 105 (26.0%) oftentimes takes their launch between 4-5days/week and 6-7days/week, respectively.

On the main place the students take their dinner in a typical week, 371 (89.8%) take their dinner at home and the rest, 42 (10.2%) take their dinner outside. The majority 230 (55.7%) oftentimes take their dinner between 6-7days/week.

The result of the often times the students use selected foods in a week were presented in table 4. On dairy foods (Skim milk, Whole milk, Yogurt, Ice cream, Cheese, Margarine, Butter) majority 189 (45.8%) take it/them 2-3days/week. More than half proportion of the students 214 (51.8%) take Fruits such as apples, pears, watermelon, oranges, orange-juice, bananas, other fruits, fresh frozen or canned 2-3days/week. Majority, 229 (55.4%) take Vegetables such as Tomatoes, Carrots, Broccoli, Cabbage, Cauliflower, Ugu leaf etc. 2-3days/week. On meat, fish or poultry, 172 (41.6%) take them 2-3days/week. Greater proportion 147 (35.6%) take starchy staples such as Cereals, Roots and Tubers, 2-3days/week. More than half proportion of the students 255 (51.7%) take legumes and nuts such as Beans, Peanut, Cashew Nut etc. It is interesting that greater proportion 338 (81.8%) of the students does not take beverage like beer and 304 (73.6%) do not take beverage like wine and again 331 (80.1%) do not take beverages such as liquor, whiskey and gin. More so, 244 (59.1%) do not take beverages such as Hawaiian punch, lemonade, or other fruit drinks.

**Table 4:**
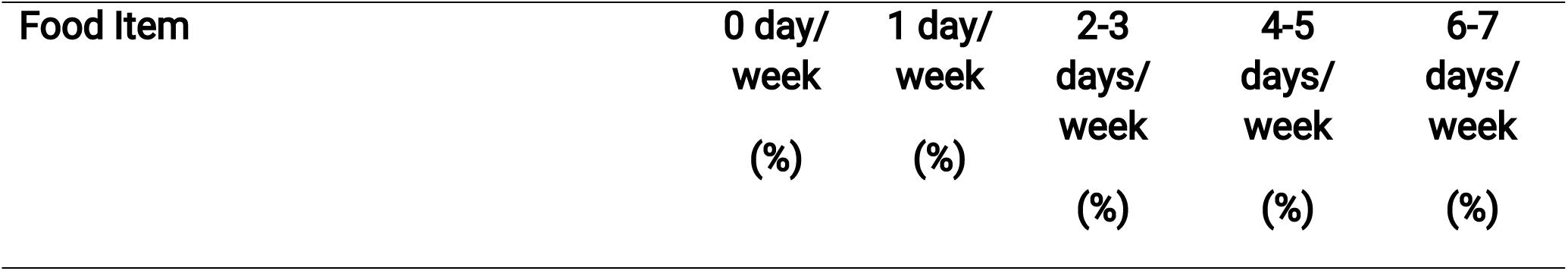
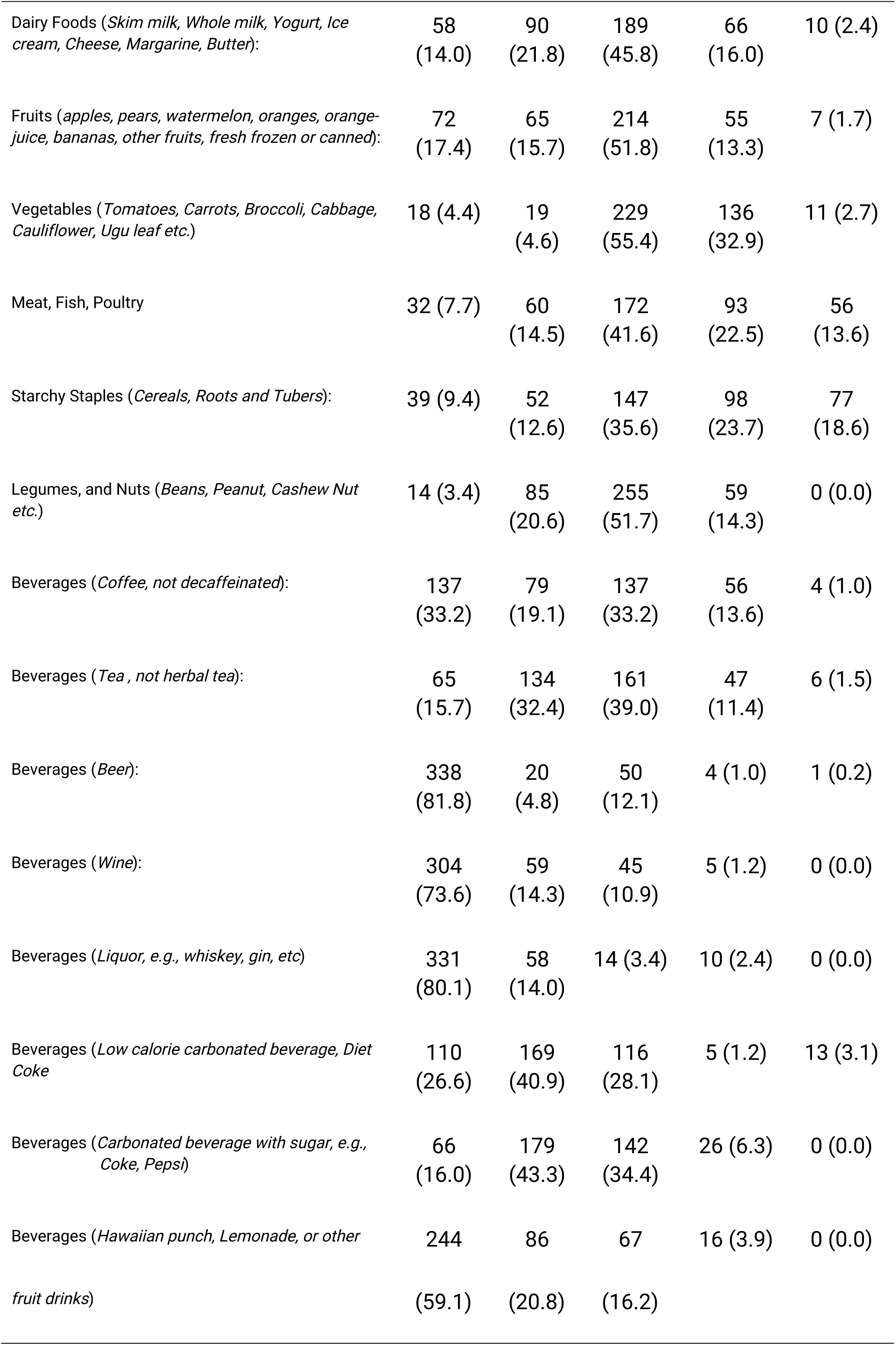
Often Times the Students use the Selected Foods in a Week in the Past 12Months.

The determinants of dietary patterns among the students were shown in table 5 above. Out of the nine (9) items listed, only two (2) of them were accepted as determinants of the dietary pattern since the mean score was more than the cut-off point of 3.0. The items accepted with a mean score of more than 3.0 include; “I eat certain food because of time constraints” (mean score=3.09), “It is important that the food I eat can be bought in the market or restaurant close to where I live” (mean score=3.09).

**Table 5:** Determinants of Dietary Pattern among the Students.

| Items | SD<br>(%) | D<br>(%) | Ne<br>(%) | A<br>(%) | SA<br>(%) | Sum | $\bar{x} \pm SD$ | Decision |
| --- | --- | --- | --- | --- | --- | --- | --- | --- |
| The main reason for choosing a food is its low price | 159<br>(38.5) | 106<br>(25.7) | 103<br>(24.9) | 28<br>(6.8) | 17<br>(4.1) | 877 | 2.12±1.12 | Rejected |
| The cost of food determines the number of times I eat daily | 105<br>(25.4) | 65<br>(15.6) | 169<br>(40.9) | 57<br>(13.8) | 17<br>(4.1) | 1055 | 2.55±1.13 | Rejected |
| I choose food I consume because it is easy and convenient to prepare | 66<br>(16.0) | 86<br>(20.8) | 14<br>(30.0) | 121<br>(29.3) | 16<br>(3.9) | 1174 | 2.84±1.13 | Rejected |
| I eat certain food because of time constraints | 63<br>(15.3) | 72<br>(17.4) | 90<br>(21.8) | 124<br>(30.0) | 64<br>(15.5) | 1293 | 3.13±1.30 | Accepted |
| It is important that the food I eat can be bought in the market or restaurant close to where I live | 85<br>(20.6) | 54<br>(13.1) | 55<br>(13.3) | 178<br>(43.1) | 41<br>(9.9) | 1275 | 3.09±1.33 | Accepted |
| The food I eat is highly influenced by my family/friends/colleagues' dietary pattern | 114<br>(27.6) | 53<br>(12.8) | 153<br>(39.0) | 33<br>(8.0) | 60<br>(14.5) | 1111 | 2.69±1.34 | Rejected |
| My religion affects the food I eat | 235<br>(56.9) | 92<br>(22.3) | 70<br>(16.9) | 16<br>(3.9) | 0<br>(0.0) | 693 | 1.68±0.89 | Rejected |
| My culture affects the food I eat | 194<br>(47.0) | 93<br>(22.5) | 82<br>(19.9) | 34<br>(8.2) | 10<br>(2.4) | 812 | 1.97±1.10 | Rejected |
| I usually eat food that is trendy | 261<br>(63.2) | 53<br>(12.8) | 91<br>(22.0) | 8<br>(1.9) | 0<br>(0.0) | 680 | 1.65±0.95 | Rejected |
| **Mean Determinant** |  |  |  |  |  | **997** | **2.41±0.52** | **Rejected** |

Others that were near acceptance with mean score more than 2.50 but were rejected include; “I choose food I consume because it is easy and convenient to prepare” (Mean score=2.84), “The cost of food determines the number of times I eat daily” (Mean score=2.55), and “The food I eat is highly influenced by my family/friends/colleagues’ dietary pattern” (Mean score=2.69).

Generally, with a weighted mean less than 3.0 of the items that measure the determinant of dietary pattern among the students (mean score=2.41, SD=0.52),it means that even though some items were accepted, there were total rejection of items that measures the determinants of dietary pattern among the students.

On the association between the socio-demographic characteristics of the students and the determinants of their dietary pattern as presented in table 6. There is no significant influence of the age group of the students on the determinants of their dietary pattern (F = 0.731; P = 0.534 > 0.05). Again, there is no significant association between gender (t = 0.585; P = 0.559 >0.05), marital status (t = 1.282; P = 0.200 > 0.05), and the ethnicity (t = 0.802; P = 0.423 > 0.0) of the students and the determinants of their dietary pattern.

**Table 6:** Association of Socio-Demographic Characteristics and Determinants of Dietary Pattern among the Students.

| Characteristics | n | Mean | SD | F/t | P-Value |
| --- | --- | --- | --- | --- | --- |
| Age Group (Years) |  |  |  |  |  |
| 15-19 | 95 | 2.46 | 0.47 | 0.731 | 0.534 |
| 20-24 | 228 | 2.42 | 0.52 |  |  |
| 25-29 | 64 | 2.34 | 0.62 |  |  |
| 30 & above | 26 | 2.36 | 0.48 |  |  |
| Gender |  |  |  |  |  |
| Male | 182 | 2.40 | 0.54 | 0.585 | 0.559 |
| Female | 231 | 2.43 | 0.51 |  |  |
| Marital Status |  |  |  |  |  |
| Single | 401 | 2.42 | 0.52 | 1.282 | 0.200 |
| Married | 12 | 2.22 | 0.55 |  |  |
| Religion |  |  |  |  |  |
| Christian | 400 | 2.42 | 0.52 | 2.486 | 0.013 |
| Others (Islam, Atheist, Traditional) | 13 | 2.06 | 0.44 |  |  |
| Ethnicity |  |  |  |  |  |
| Igbo | 392 | 2.41 | 0.51 | 0.802 | 0.423 |
| Others (Yoruba, Hausa) | 21 | 2.50 | 0.73 |  |  |
| **Level of Study** |  |  |  |  |  |
| 200L | 134 | 2.51 | 0.53 | 2.702 | 0.030 |
| 300L | 88 | 2.46 | 0.46 |  |  |
| 400L | 84 | 2.33 | 0.50 |  |  |
| 500L | 72 | 2.34 | 0.60 |  |  |
| 600L | 35 | 2.30 | 0.49 |  |  |
| **Monthly Allowance (Naira)** |  |  |  |  |  |
| <10,000 | 149 | 2.42 | 0.49 | 3.705 | 0.006 |
| 10,000-20,000 | 127 | 2.52 | 0.56 |  |  |
| 21,000-30,000 | 44 | 2.35 | 0.51 |  |  |
| 31,000-40,000 | 61 | 2.21 | 0.36 |  |  |
| 41,000-50,000 | 32 | 2.43 | 0.69 |  |  |

However, there is a significant association between the religion of the participants and the determinants of their dietary pattern (t = 2.486; P = 0.013 < 0.05). This implies that the Christians were considerate of their dietary pattern with mean = 2.42 more than others.

Again, there is a significant association between the level of study of the participants and the determinants of their dietary pattern (F = 2.702; P = 0.030 < 0.05). This implies that the students in their 200levels were considerate of their dietary pattern with mean = 2.51 more than others.

More so, there is a significant association between the monthly allowances received by the students and the determinants of their dietary pattern (F = 3.705; P = 0.006 < 0.05). This implies that the students who receive between 10,000 to 20,000 with mean = 2.52and those that receive between 41,000 to 50,000 with mean = 2.43 were considerate of their dietary pattern more than others.

## CHAPTER FIVE

### 5.0 DISCUSSION

The survey attempted to clearly answer the research questions on the dietary pattern among medical students of Ebonyi State University in Abakaliki, Nigeria.

Among the respondents, 182 (44.1%) are males, 231 (55.9%) are females. Also, the respondents 95 (23%) are aged 15 to 19 years, 228 (55.2%) are aged 20 to 24 years, 64 (15.5%) are aged 25 to 29 years while 26 (6.3%) are aged 30 years and above.401 (97.1%) are single 12 (2.9%) are married. Most 400 (96.9%))are Christians.392 (94.9%) are Igbo by tribe while 8 (4.4%) are from other tribes (Yoruba, Hausa).On the level of study among the respondents, 134 (32.4%) are in 200 level, 88 (21.3%) are in 300 level, 84 (20.3%) are in 400level, 72 (17.4%) are in 500 level, 85 (8.5%) are in 600 level. Most receive monthly allowance of less than #10,000, 149 (36.1%).

In our study to assess the attitude of the medical students towards diet, only about 133 (32.2%) take most of all responsibilities for food shopping,132 (32%) take most of all responsibilities for planning of meals while 186 (45%) take most of all responsibilities for preparing meals. This shows that a greater proportion of the students take about half, little or no responsibilities for food shopping, planning of meals and preparing of meals.

On the pattern of diet among the medical students, it is seen that 334 (80.9%), 274(66.3%)and 371(89.8%) take their breakfast, lunch, and dinner at home respectively. This implies that greater proportion of the students of the students take their breakfast, lunch and dinner at home while a lesser proportion of the students eat outside or do not eat at all. However, Out of the 370 that take their breakfast either at home or outside, 123 (33.2%) and 136 (36.8%) oftentimes take their breakfast between 4-5days/week and 6-7days/week, respectively. Also, out of the 404 that take their lunch either at home or outside,177 (43.8%) and 105 (26.0%) oftentimes takes their lunch between 4-5days/week and 6-7days/week, respectively. This shows that greater proportion of the students often times take their breakfast, lunch and dinner respectively for 4 to 7 days in a week.

This study also ascertained how often the students use selected foods in a week for the past 12 months and it is found from the study that majority (45.8%) take diary food 2 to 3 days/week. More than half proportion of the students (51.8%) take fruits such as apples, pears, watermelon etc 2 to 3 days/week. Majority (55.4%) take vegetables such as tomatoes, carrots, broccoli, etc 2 to 3 days/week. On meat, fish, or poultry (41.6%) take them 2 to 3 days/week.Greater proportion (35.6%) take starchy stapples such as cereals, roots, and tubers,2 to 3 days/week. Meanwhile, a proportion (81.8%) of the students do not take beverage like beer, 73.6% do not take beverages like wine and 80.1% do not take beverages such as liquor, whiskey, and gin. More so,59.1% of the respondents do not take beverages such as hawaiian punch, lemonade, or other fruit drinks. This explanation means that the dietary diversity of the medical students is poor/low.

Out of the nine items that were studied as the determinants of dietary pattern among medical students, only two items were accepted because they met the cut-off point (mean score > or = 3.0). Consequently, the major determinants of dietary pattern among medical students of Ebonyi State University include; time constraints(with a mean score = 3.09) and proximity of the needed food(with a mean score = 3.09). However, other factors that were seen to have significant association with the determinants of dietary pattern include; religion with a P value = 0.013 < 0.05, the level of study with a P value = 0.030 < 0.05, and the monthly allowance with a P value = 0.006 < 0.05. This implies that religion, level of study, and monthly allowance significantly affected the dietary pattern of the medical students.

## CHAPTER SIX CONCLUSION AND RECOMMENDATIONS

### 6.0 CONCLUSION

Diet is an important and also modifiable life-style determinant of human health. For students, dietary pattern can also influence their academic performance. It can be deduced from the findings from this study that majority of the medical students have poor attitude towards diet and low/poor dietary diversity owing to the number of times (2 to 3 days/week) majority of the students use the selected food in a week in the past 12 months.It was also observed from this study that time constraints and proximity of the needed food are the two major determinants of dietary pattern among the medical students. Other factors that have significant influence on their dietary pattern include; religion, level of study and monthly allowance.

### 6.1 RECOMMENDATIONS

Based on the findings of this study, the following recommendations are made:

Nutritional interventions should be implemented at an individual, family, community and national levels.

At individual level, behavioural change communications should be targeted towards the students to improve their attitude towards diet and also improve on their dietary diversity.

At family and community level, parents, guardians, sponsors, as well as philanthropists should be sensitized on the influence of the monthly allowance of the students on their dietary pattern.

At national level, full scholarship should be awarded to the indigent students by the government.

## Data Availability

All data produced in the present work are contained in the manuscript.

## QUESTIONNAIRE

### TITLE: ASSESSMENT OF DIETARY PATTERN AMONG MEDICAL STUDENTSOFEBONYISTATEUNIVERSITY,ABAKALIKI,NIGERIA

Dear respondent,

You are kindly requested to give your sincere responses to the following questions to ensure that analysis is without bias. Your identity is not required. You are hereby assured that any information volunteered will be treated with utmost confidentiality. The questionnaire aims to assess the Pattern of Diet among the Medical Students of Ebonyi State University.

Thank you.

### SECTION A: SOCIO-DEMOGRAPHIC CHARACTERISTICS

1. **Age (years):** (a) 15 - 19(b)20-24 (c) 25 - 29 (d) 30 - 34 (e) 35 - 39 (f) 40 - 44
2. **Gender:** Male (a) Female (b)
3. **Marital Status:** (a) Single (b) Married (c) Divorced (d) Widowed
4. **Religion:** (a) Christian (b) Islam (c) Traditional (d) Atheist
5. **Ethnic Group:** (a) Igbo (b) Yoruba (c)Hausa (d)Others, specify ()
6. **Level of Study:** (a)100L (b) 200L (c) 300L (d) 400L (e) 500L (f) 600L
7. **Monthly Allowance (in naira):**(a) <10,000 (b) 10,000 - 20,000 (c) 21,000 - 30,000 (d) 31,000 - 40,000(e) 41000-50,000

### SECTION B: ATTITUDE TOWARDS DIET

8. How much responsibility do you have for Food shopping? (a)Little or none(b) About half (c) Most or all
9. How much responsibility do you have for Planning meals? (a)Little or none(b) About half (c) Most or all
10. How much responsibility do you have for Preparing meals? (a)Little or none(b) About half (c) Most or all

### SECTION C: PATTERN OF DIET

11. Where are most of your Breakfasts in a typical week? (a) At home (b) Outside (c) Don’t eat
12. How often do you take your Breakfast in a typical week? (a)l day/ week(b) 2-3 days/week (c) 4-5 days/week(d) 6-7days/week(
13. Where are most of your Lunches prepared in a typical week? (a) At home (b) Outside (c) Don’t eat
14. How often do you take your lunch in a typical week? (a)l day/ week(b) 2-3 days/week (c) 4-5 days/week (d) 6-7days/week
15. Where are most of your Dinners prepared in a typical week? (a) At home (b) Outside(c) Don’t eat
16. How often do you take your dinner in a typical week? (a)l day/ week(b) 2-3 days/week (c) 4-5 days/week (d) 6-7days/week

#### For each food listed, indicate how often on average you use them during the past year

17. DAIRY FOODS (Skim milk, whole milk, yogurt, ice cream, cheese, margarine, butter): *(a) 0 day/week (b)l day/ week(c) 2-3 days/week (d) 4-5 days/week (e) 6-7days/week*
18. FRUITS (apples, pears, watermelon, oranges, orange juice, bananas, other fruits, fresh frozen or canned):(a) 0 day/week(b)1 day /week(c) 2-3 days/week(d) 4-5 days/week(e) 6-7 days/week
19. VEGETABLES (tomatoes, carrots, broccoli, cabbage, cauliflower, ugu leaf etc): (a) 0 day/week (b)1 day/week(c) 2-3 days/week (d) 4-5 days/week(e) 6-7 days/week
20. MEAT,FISH,POULTRY: (a)0 day/week (b)l day/week (c) 2-3 days/week(d) 4-5 days/week (e) 6-7.days/week
21. STARCHY STAPLES (cereals, roots and tubers): (a)0 day/week (b)l day/week (c) 2-3 days/week (d) 4-5 days/week (e) 6-7 days/week
22. LEGUMES. AND NUTS(beans, peanut, cashew nut etc): (a)0 day/week (b)l day/week (c) 2-3 days/week(d) 4-5 days/week (e) 6-7 days/week
23. BEVERAGES (Coffee, not decaffeinated): (a)0 day/week (b)1 day /week(c) 2-3 days/week (d) 4-5 days/week (e) 6-7 days/week
24. BEVERAGES (Tea, not herbal tea): (a)G day/week(b)1 day /week (c) 2-3 days/week (d) 4-5 days/week (e) 6-7 days/week
25. BEVERAGES (Beer): (a)0 day/week (b)l day /week (c) 2-3 days/week(d) 4-5 days/week (e) 6-7 days/week
26. BEVERAGES (Wine): (a) 0 day/week (b) 1 day /week (c) 2-3 days/week (d) 4-5 days/week (e)6-7days/week
27. BEVERAGES (Liquor, e.g., whiskey, gin, etc): (a)0 day/week (b)1 day /week (c) 2-3 days/week (d) 4-5 days/week (e) 6-7 days/week
28. BEVERAGES (Low calorie carbonated beverage, Diet Coke) (a)0 day/week(b)l day /week (c) 2-3 days/week(d) 4-5 days/week(e) 6-7 days/week
29. BEVERAGES (Carbonated beverage with sugar, e.g., Coke, Pepsi) (a)0 day/week (b)l day /week (c) 2-3 days/week (d) 4-5 days/week(e) 6-7 days/week
30. BEVERAGES (Hawaiian Punch, lemonade, or other fruit drinks) (a) 0 day/week(b)l day /week(c) 2-3 days/week(d) 4-5 days/week (e) 6-7 days/week
31. What is the average volume of water you drink in a day? (a) <1L (b) 1L (c) 2L - 3L (d) 4L (e) > 4L
32. Do you currently follow a special diet? Number of years on diet? (a)Yes (b) No
33. If yes, for how many years?(a) <5 Yrs(b) 5-10yrs (c) >10 yrs
34. If yes, what kind of diet do you follow? (Select more than one if necessary.) (a) Weight reduction (low calorie)(b) Diabetic (c) Ulcer (d) Low cholesterol/ Low fat(e) High Potassium (f) Low sodium()

### SECTION D: DETERMINANTS OF DIETARY PATTERN

35. The main reason for choosing a food is it’s low price (a)Strongly Disagreed (b)Disagreed (c)Neutral (d) Agreed (e)Strongly Agreed()
36. The cost of food determines the number of times I eat daily (a) Strongly Disagreed (b)Disagreed (c)Neutral (d)Agreed (e)Strongly Agreed
37. I choose food I consume because it is easy and convenient to prepare (a) Strongly Disagreed (b)Disagreed (e)Neutral (d) Agreed (e)Strongly Agreed
38. I eat certain food because of time constraints (a)Strongly Disagreed (b)Disagreed (c)Neutral (d)Agreed (e)Strongly Agreed
39. It is important that the food i eat can be bought in the market or restaurant close to where I live(a)Strongly Disagreed (b)Disagreed (c) Neutral (d)Agreed (e)Strongly Agreed
40. The food I eat is highly influenced by my family/friends/colleagues’ dietary pattern (a)Strongly Disagreed (b)Disagreed (c)Neutral (d)Agreed (e)Strongly Agreed
41. My religion affect the food I eat (a)Strongly Disagreed (b)Disagreed (c)Neutral (d)Agreed (e)Strongly Agreed
42. My culture affect the food I eat (a)Strongly Disagreed (b)Disagreed (c)Neutral (d)Agreed (e)Strongly Agreed
43. I usually eat food that is trendy (a) Strongly Disagreed (b) Disagreed (c)Neutral (d)Agreed (e)Strongly Disagreed

